# Synthesizing evidence regarding community-based volunteer facilitated programs supporting integrated care transitions from hospital to home: A scoping review protocol

**DOI:** 10.1101/2021.02.20.21251514

**Authors:** Michelle L. A. Nelson, Alana Armas, Rachel Thombs, Hardeep Singh, Joseph Fulton, Heather Cunningham, Sarah Munce, Sander L. Hitzig, Janet Prvu Bettger

## Abstract

**Background:** Given the risks inherent in care transitions, it is imperative that patients discharged from hospital to home receive the integrated care services necessary to ensure that the transition is successful. Despite efforts by the health care sector to develop health system solutions to improve transitions, problems persist. Research on transitional support has predominantly focused on services delivered by health care professionals; the evidence for services provided by lay navigators or volunteers in supporting people transitioning from hospital to home has not been synthesized to guide practice, policy or future research.

**Aim:** This is a protocol for a scoping review that will examine the role and contributions of voluntary sector personnel and services to support transitions from hospital to home.

**Methods:** Using the well-established scoping review methodology outlined by Arksey and O’Malley, a six-stage study is outlined (1) identifying the research question, (2) identifying relevant studies, (3) selecting studies, (4) charting the data, (5) collating, summarizing and reporting the results, and (6) providing consultation. The search strategy, designed by an information scientist, is applied to ten databases reflecting empirical and grey literature sources. A two-stage screening process will be used to determine eligibility of articles. To be included in the review, articles must report on a community-based program that engages volunteers in the provisions of services that support adults transitioning from hospital to home. All articles will be independently assessed for eligibility, and data from eligible articles will be abstracted and charted using a standardized form. Extracted data will be analyzed using narrative and descriptive analyses. Research ethics approval is not required for this scoping review.

**Discussion:** This scoping review will map the available literature focused on the contributions of voluntary sector personnel and services to support transitions from hospital to home.

## Background

Transitioning from hospital to home is well recognized to be a time of increased vulnerability [1-9]. People who may already be vulnerable, such as the elderly, immigrants, and individuals with complex chronic conditions, are even more so during transitions and may require additional support [9-13]. By definition, a transition of care is “a set of actions designed to ensure the coordination and continuity of health care as patients transfer between different locations or different levels of care within the same location” [14]. Irrespective of the care trajectory a person follows, they will undergo at least one transition during their time as an inpatient, either from one care setting or level of care to another, or through discharge back home [14-16]. Transitions are inherently complex, requiring well-coordinated efforts by many individuals across multiple settings [14, 17]. Ideally, transitions involve comprehensive care plans that include community follow-up and timely information exchange between hospital and community practitioners about a patient’s treatment goals, preferences and their health and clinical status [17-20]. However, in reality, there is growing evidence of failures in provider communication and fragmented care during care transitions [19-21]. Fragmentation can lead to adverse events, including medication errors, readmissions, decreased patient satisfaction, further morbidity and even mortality [22-27]. These issues can be exacerbated in communities where resources and services are lacking, transportation limitations exist, providers are limited or wait times are increased [26-31]. It is estimated that 20% of patients are re-hospitalized within 30 days post hospital discharge; which can be quite costly to a healthcare system [6]. Given the risks associated with transitions [1-9], it is imperative that those discharged home from hospital receive the care and assistance necessary to ensure that their care transition is safe and successful. Despite efforts of the health care sector to develop health system solutions to improve transitions, problems persist [32, 33].

Efforts to address issues related to transitions of care are occurring at all levels in the health and social care sectors. Increasingly complex patient populations [34], new reimbursement models [35], shortened lengths of hospital stay [36] and pressures to reduce costs while improving care quality and patient experience have driven policy makers to seek innovative solutions. Spurred by health reforms, health system leaders have turned to partnerships with community-based organizations (CBOs) to improve patient care [37]. Non-government CBOs are situated in the communities they serve, have a deep understanding of members’ needs, and are in a trusted position to help [38]. Partnerships with and between non-government CBOs are a recognized way to expand the breadth and quality of health and social services [39-41]. It has also been suggested that the public sector needs these partnerships as the knowledge, skill and innovation inherent to non-government CBOs are indispensable to running a welfare state [42]. The Health Systems Learning Group, comprised of 43 health care organizations from across the USA, noted that “as hospitals and health systems struggle under the weight of uncompensated care, emergency department (ED) overuse, and readmissions—the greater portion directly attributable to spiraling chronic disease—the case for transformative community partnerships becomes increasingly clear” (p.65) [43]. For example, the ‘*Memphis Model’* a partnership between more than 600 congregations and Methodist Le Bonheur Healthcare to support congregants post discharge through the collaborative efforts of hospital employed navigators and trusted volunteers from each parish [44]. This model is seen to create synergy between different types of organizations focused on health and wellbeing, ultimately being of value to patients, congregations and community more broadly [45]. However, the role and contributions of a key community partner – non-government organizations (e.g., voluntary sector, charitable, third sector) - in improving patient experience and system efficiency are still often unaccounted for, or under-recognized [45].

While these partnerships between health system leaders and non-government CBOs represent engagement at the macro and meso levels, a variety of support roles at the micro level are another way that volunteers and the voluntary sector are contributing towards successful transitions. Lay navigators are one such intervention; individuals recruited from the community who receive specialized training to assist patients in navigating complex health and social care systems. Post discharge, lay navigators are involved to help with maintaining health service engagement through assistance with scheduled appointments and referrals, providing accompaniment to appointments, communicating with relevant agencies and organizations, and assisting with paperwork and forms [46-48]. In addition, lay navigators also signpost and help adults with chronic illness living in the community build connections with their community with the intent to improve quality of life, and develop independence and engagement [49].

Recognizing the potential value of voluntary sector services in supporting patient care, organizations such as The Kings Fund [50], the Institute for HealthCare Improvement [51], and The Beryl Institute [52] have advocated for more purposeful engagement of non-government CBOs in the organization and delivery of health care. Research on the engagement of volunteers, lay navigators and partnerships with non-government CBOs as forms of integrated care in supporting care transitions is emerging, but the evidence base is unclear in comparison to transitional support services delivered by health care professionals [53-55]. To address this knowledge gap, this scoping review will map the available literature focused on the contributions of voluntary sector personnel and services to support transitions from hospital to home.

## Methods/Design

Published literature on community-based volunteer supported transition programs is likely to appear in a variety of sources and vary in methodological approaches and formats. Using the Arksey and O’Malley Framework [56] and the PRISMA-Scoping Review reporting guidelines will allow us to explore the broad topic and make use of literature across study designs and in both peer reviewed and grey literature. The study will be completed over six stages: (1) identifying the research question, (2) identifying relevant studies, (3) selecting studies, (4) charting the data, (5) collating, summarizing and reporting the results, and (6) providing consultation based on the findings. Though it does not entail quality assessment, scoping reviews are considered a rigorous and systematic approach to knowledge synthesis. Ethics Review Board approval is not required for the conduct of this study.

### Identifying the research question

Members of this research team had previously convened experts to identify high priority research questions specific to rehabilitation for stroke patients with multiple concurrent health and social issues [57]. During this consensus meeting, several questions arose about the role and potential of ‘non health sector’ organizations [57] which supported the development of the research question for this scoping review. Based on knowledge users’ feedback, the study question was not limited to the stroke population, since programs developed for other patient populations may be transferable. This review will answer the following research question: *What is the extent and nature of the literature describing lay navigators’, volunteers’ and non-government CBOs’ engagement in supporting adults in the transition from hospital to home?*

#### Study objectives include

1. To determine how volunteers of non-government CBOs have been engaged to support adults being discharged home from the hospital.
2. To identify key program characteristics, facilitators, barriers, and outcomes of relevance specific to transitions and community reintegration practices delivered by volunteers of non-government CBOs.
3. To ascertain stakeholder perspectives regarding transitions and community reintegration services delivered by volunteers of non-government CBOs.
4. To identify relevant knowledge gaps that can support the development of a research program focused on transitions and community reintegration with volunteers as a key service provider.

### Identifying relevant studies

To identify relevant peer-reviewed studies the research team, along with an information scientist (HC), developed a comprehensive search strategy. The initial search strategy was generated for OVID Medline and peer reviewed by information science librarians, after which the information scientist adopted the search to the MeSH terms and concepts for the remaining databases. Keywords included volunteer*, voluntary health agencies, faith-based organizations, post discharge, transition*, after-care, post-hospital, self-help, peer support, patient discharge, community reintegration, unpaid, home and community care. Due to the nature of the research question, the team recognized that potentially relevant literature would be identified in a wide variety of databases. After careful consideration, nine additional databases were selected for the review: EMBASE, PsycInfo, Joanna Briggs (JBI), Social Work Abstracts, Sociological Abstracts, CINAHL, Cochrane Reviews, Ageline, and Scopus. All searches will be limited to records published from 2000 to present. The preliminary database search has identified 19, 720 records on OVID Medline. Reference lists and bibliographies of the identified articles will also be searched for citations not identified by the database search. A gray literature search will be conducted to identify any non-indexed literature of relevance. The gray literature search will focus on organizational reports, conference proceedings, theses and dissertations. All literature searches will be conducted by the experienced Information Scientist on the study team (HC). Finally, other global experts in transitions will be consulted in a ‘desk drawer’ search strategy to ensure that all relevant citations are obtained. The studies included in the review will be amalgamated and stored using reference management software package, EndNote, to ensure there are no duplicates in the database.

### Study selection

Once all identified records have been extracted from all databases and duplicates have been removed, the data will be uploaded into knowledge synthesis management platform, Covidence™, for screening and abstraction. We will include published and unpublished literature reporting any quantitative, qualitative, mixed or multi methods research, including both comparative (e.g., randomized, controlled, cohort, quasi-experimental) and non-comparative (e.g., survey, narrative, audit) methods, educational materials and reports related to the topic of treatment goal setting for complex patients. We will include all articles that meet the following criteria: (1) articles specifically describe community-based programs (non-government, charitable, voluntary sector) that support hospital discharge and return to home, (2) one component of the program is delivered by volunteers as the program workforce, (3) programs focused on adult populations. Programs and services offered to children and adolescents are different from programs for adults, as they incorporate guardians and address needs that are specific to those under the age of 18. These differences constitute a separate review of the literature that goes beyond the scope of the current review and therefore will be excluded.

The inclusion and exclusion criteria will be tested on a randomly selected group of studies. Once the final inclusion and exclusion criteria have been determined, the team will pull another random sample of identified articles to screen and test inter-rater reliability, using the Kappa coefficient. We will continue to test inter-rater reliability until a Kappa rate of 0.85 is reached. Title and abstract screening will be conducted in duplicate on all identified articles. Based on the inclusion and exclusion criteria, the reviewers will categorize the articles as ‘Yes’, ‘No’ or ‘Maybe’; all ‘Yes’ and ‘Maybe’ articles will be included for full-text screening. Any discrepancies will be reviewed and resolved by the senior research team members and experts in this field. Inter-rater reliability will be continuously tested throughout the title and abstract screen to ensure a high rating is maintained. This will also give the research team multiple opportunities to discuss and resolve discrepancies. For full-text screening, the reviewers will categorize the articles as either ‘Yes’ or ‘No’, and any uncertainties will be discussed by the team with discrepancies adjudicated by a senior member of the research team.

### Charting the data

A copy of each article/document will be obtained, reviewed and charted by two research team members. Data abstraction will be completed by specified research team members for all articles (i.e., two researchers per article, with adjudication by a third researcher) using an abstraction form that will be pilot tested before use. During pilot testing abstraction criteria may be modified for full abstraction of the included articles. Preliminary data abstraction elements will include:

- Researcher performing data abstraction and date of data abstraction
- Identification features of the article (record number, author, year)
- Type of publication (published or unpublished)
- Study Design
- Program Details:
  - How “transition support” is conceptualized/defined;
  - Organizational Profile (size, location);
  - Program aims and objectives;
  - Services provided and delivery mechanisms (e.g., client characteristics, eligibility, duration, referral processes, assessments);
  - Service administration and demographics (e.g., funding, staffing mix, volunteer characteristics and requirements); and
  - Evaluation procedures and outcomes.

During abstraction, any discrepancies will be reviewed and resolved by a senior member of the research team.

### Collating, summarizing and reporting the results

A scoping review is designed to provide an overview of the extent and nature of a body of literature. To do this, we will employ three reporting and presentation strategies: (a) a modified Preferred Reporting Items for Systematic Reviews and Meta-Analysis (PRISMA-SR) (b) a basic numerical account of the amount, type, and distribution of the studies included in the review, and (c) a thematic analysis and map of included literature.

The specific reporting format and products will be determined by the results and needs of our knowledge users. We anticipate the production of tables and charts will organize and depict references according to publication type, program details (e.g., location, funding, outcomes, staffing), and client characteristics (e.g., age, condition, ethnicity). The anticipated breadth of the literature spanning conditions, setting, age, and location makes it challenging to adopt an existing conceptual framework to comprehensively map the literature. As a result, we anticipate that we will have to develop a framework in order to summarize and present the results of this review.

### Consultation and Dissemination

Throughout data extraction and analysis of the included studies, we will consult with members of an existing international special interest group focused on the voluntary sector in integrated care. This group will provide insight and feedback on study findings, help with dissemination of the results, and engage in the development of future research proposals. Traditional end of grant dissemination activities, including peer reviewed publications and academic presentations at local, national and international conferences are planned.

## Discussion

This scoping review will generate a high-quality synthesis of knowledge regarding the role and contributions of voluntary sector personnel and services to support transitions from hospital to home. Although volunteer service and engagement can be valuable to improving the quality of transitions, this topic remains understudied.

There are several noteworthy strengths of the proposed scoping review. First, the application of a rigorous, well-established methodological framework will ensure production of a high-quality review [58]. Second, we have included rich details of our methodology to enhance clarity and transparency and demonstrate procedural and methodological rigour [58]. Third, we will ensure reliability between screeners with a higher inter-rater reliability. Inter-rater reliability is an important variable that will ensure the records screened are “correct representations of the variables measured” [59]. Fourth, our screening team includes content and methodological experts; according to best practice guidelines for screening, this is key to conducting a high-quality review [60]. Finally, given the broad nature of the research question, we will conduct a comprehensive search on 10 disciplinary and cross-disciplinary databases [58]. This will ensure that we will maximize our coverage of all possible records that meet review inclusion criteria [61]. Finally, the inclusion of gray literature further strengthens our review by reducing publication bias and enhancing comprehensiveness of the findings [62].

However, some study limitations are anticipated in this review. First, the synthesis will be limited to articles published in English. Despite the risk of missing relevant programs reported in languages other than English, this limitation was imposed to facilitate a timely review given the high number of citations that had been returned from our preliminary database search (i.e., 19, 720 citations) and the high volume of citations anticipated from additional databases. Second, if a sufficient inter-rater reliability is reached, we may employ a single screener approach. A single screening is one strategy to facilitate timely study conduct. However, we acknowledge that this may lead to some relevant articles/literature being missed. To compensate for this, if single screening methods are employed, hand searches of the reference lists of the included articles will be conducted. Third, despite our comprehensive search strategy, the lack of standardized terminology for volunteer/lay navigators in the literature creates a possibility that relevant articles may be missed. However, hand searches of reference lists of the included studies may minimize this limitation.

## Data Availability

Data is not available.

## List of abbreviations

CBO: Community-based organization

## Declarations

### Ethics approval and consent to participate

Not applicable.

### Consent for publication

Not applicable.

### Availability of data and materials

Not applicable.

### Authors’ Contributions

MN conceptualized and designed the study and wrote and critically revised the protocol and manuscript. AA and RT wrote and critically revised the protocol and manuscript and collaborated with the information scientist on the search strategies. HS, SH, JPB and SM critically revised the protocol and manuscript. JF collaborated with the information scientist on the grey literature search strategy and revised the protocol and manuscript. As the information scientist, HC devised the peer-reviewed and grey literature search strategies in collaboration with the team.

## Acknowledgements

Not applicable.

## References

1. Neiterman E, Wodchis WP, Bourgeault IL: Experiences of older adults in transition from hospital to community. Can J Aging 2015, 34(1):90–99.

2. Naylor MD: Transitional care for older adults: a cost-effective model. LDI Issue Brief 2004, 9(6):1–4.

3. Krumholz HM: Post-hospital syndrome--an acquired, transient condition of generalized risk. N Engl J Med 2013, 368(2):100–102.

4. Foust JB, Naylor MD, Bixby MB, Ratcliffe SJ: Medication problems occurring at hospital discharge among older adults with heart failure. Res Gerontol Nurs 2012, 5(1):25–33.

5. Andreasen J, Lund H, Aadahl M, Sørensen EE: The experience of daily life of acutely admitted frail elderly patients one week after discharge from the hospital. Int J Qual Stud Health Well-being 2015, 10:27370.

6. Jencks SF, Williams MV, Coleman EA: Rehospitalizations among patients in the Medicare fee-for-service program. N Engl J Med 2009, 360(14):1418–1428.

7. Mesquita ET, Cruz LN, Mariano BM, Jorge AJL: Post-Hospital Syndrome: A New Challenge in Cardiovascular Practice. Arq Bras Cardiol 2015, 105(5):540–544.

8. van Seben R, Reichardt LA, Essink DR, van Munster BC, Bosch JA, Buurman BM: “I Feel Worn Out, as if I Neglected Myself”: Older Patients’ Perspectives on Post-hospital Symptoms After Acute Hospitalization. Gerontologist 2019, 59(2):315–326.

9. Hestevik CH, Molin M, Debesay J, Bergland A, Bye A: Older persons’ experiences of adapting to daily life at home after hospital discharge: a qualitative metasummary. BMC Health Serv Res 2019, 19(1):224.

10. Parekh AK, Goodman RA, Gordon C, Koh HK, Conditions HHSIWoMC: Managing multiple chronic conditions: a strategic framework for improving health outcomes and quality of life. Public Health Rep 2011, 126(4):460–471.

11. Baxter R, Shannon R, Murray J, O’Hara JK, Sheard L, Cracknell A, Lawton R: Delivering exceptionally safe transitions of care to older people: a qualitative study of multidisciplinary staff perspectives. BMC Health Serv Res 2020, 20(1):780.

12. Rustad EC, Furnes B, Cronfalk BS, Dysvik E: Older patients’ experiences during care transition. Patient Prefer Adherence 2016, 10:769–779.

13. Tomlinson J, Cheong VL, Fylan B, Silcock J, Smith H, Karban K, Blenkinsopp A: Successful care transitions for older people: a systematic review and meta-analysis of the effects of interventions that support medication continuity. Age Ageing 2020, 49(4):558–569.

14. Coleman EA, Boult C: Improving the quality of transitional care for persons with complex care needs. J Am Geriatr Soc 2003, 51(4):556–557.

15. Naylor M, Keating SA: Transitional care. Am J Nurs 2008, 108(9 Suppl):58–63.

16. Coleman EA, Smith JD, Frank JC, Min SJ, Parry C, Kramer AM: Preparing patients and caregivers to participate in care delivered across settings: the Care Transitions Intervention. J Am Geriatr Soc 2004, 52(11):1817–1825.

17. Waring J, Marshall F, Bishop S, Sahota O, Walker M, Currie G, Fisher R, Avery T: Health Services and Delivery Research. In: An ethnographic study of knowledge sharing across the boundaries between care processes, services and organisations: the contributions to ‘safe’ hospital discharge. edn. Southampton (UK): NIHR Journals Library; 2014.

18. Cohen MD, Hilligoss PB: The published literature on handoffs in hospitals: deficiencies identified in an extensive review. Qual Saf Health Care 2010, 19(6):493–497.

19. Abrashkin K, Cho H, Torgalkar S, Markoff B: Improving Transitions of Care From Hospital to Home: What Works? Mt Sinai J Med 2012, 79:535–544.

20. Clarke JL, Bourn S, Skoufalos A, Beck EH, Castillo DJ: An Innovative Approach to Health Care Delivery for Patients with Chronic Conditions. Pop Health Manag 2016, 20(1):23–30.

21. Abraham J, Kannampallil T, Patel B, Almoosa K, Patel VL: Ensuring patient safety in care transitions: an empirical evaluation of a Handoff Intervention Tool. AMIA Annu Symp Proc 2012, 2012:17–26.

22. Laugaland K, Aase K, Barach P: Addressing Risk Factors for Transitional Care of the Elderly – Literature review. 2011.

23. Riesenberg LA, Leitzsch J, Cunningham JM: Nursing handoffs: a systematic review of the literature. Am J Nurs 2010, 110(4):24-34; quiz 35-26.

24. Kripalani S, LeFevre F, Phillips CO, Williams MV, Basaviah P, Baker DW: Deficits in communication and information transfer between hospital-based and primary care physicians: implications for patient safety and continuity of care. JAMA 2007, 297(8):831–841.

25. Coleman EA: Falling through the cracks: challenges and opportunities for improving transitional care for persons with continuous complex care needs. J Am Geriatr Soc 2003, 51(4):549–555.

26. Danzl MM, Harrison A, Hunter EG, Kuperstein J, Sylvia V, Maddy K, Campbell S: “A Lot of Things Passed Me by”: Rural Stroke Survivors’ and Caregivers’ Experience of Receiving Education From Health Care Providers. J Rural Health 2016, 32(1):13–24.

27. Lustig D, Weems GH, Strauser D: Rehabilitation service patterns: A rural/urban comparison of success factors. J Rehabil 2004, 70:13–19.

28. Institute of Medicine Committee on Quality of Health Care in A. In: Crossing the Quality Chasm: A New Health System for the 21st Century. edn. Washington (DC): National Academies Press (US); 2001.

29. Deber RB: Health care reform: lessons from Canada. Am J Public Health 2003, 93(1):20–24.

30. Soril LJJ, Adams T, Phipps-Taylor M, Winblad U, Clement F: Is Canadian Healthcare Affordable? A Comparative Analysis of the Canadian Healthcare System from 2004 to 2014. Healthc Policy 2017, 13(1):43–58.

31. MacNeil M, Koch M, Kuspinar A, Juzwishin D, Lehoux P, Stolee P: Enabling health technology innovation in Canada: Barriers and facilitators in policy and regulatory processes. Health Policy 2019, 123(2):203–214.

32. Rochester C, Pincus K, Patel R, Reitz S: The Current Landscape of Transitions of Care Practice Models: A Scoping Review. Pharmacotherapy 2016, 36:117–133.

33. Markiewicz O, Lavelle M, Lorencatto F, Judah G, Ashrafian H, Darzi A: Threats to safe transitions from hospital to home: A consensus study in North West London primary care. Br J Gen Pract 2019, 70:bjgp19X707105.

34. Kingston A, Comas-Herrera A, Jagger C: Forecasting the care needs of the older population in England over the next 20 years: estimates from the Population Ageing and Care Simulation (PACSim) modelling study. Lancet Public Health 2018, 3(9):e447–e455.

35. Chapin RK, Chandran D, Sergeant JF, Koenig TL: Hospital to community transitions for adults: discharge planners and community service providers’ perspectives. Soc Work Health Care 2014, 53(4):311–329.

36. Bueno H, Ross JS, Wang Y, Chen J, Vidán MT, Normand S-LT, Curtis JP, Drye EE, Lichtman JH, Keenan PS et al: Trends in length of stay and short-term outcomes among Medicare patients hospitalized for heart failure, 1993-2006. JAMA 2010, 303(21):2141–2147.

37. Garrett D, Hwang A: CBOs and state medicaid programs: A key partnership for patient-centered care. Generations 2018, 42:19–23.

38. Dickinson H, Allen K, Alcock P, Macmillan R, Glasby J: The Role of the Third Sector in Delivering Social Care. 2012.

39. Brinkerhoff D, Brinkerhoff J: Public-private partnerships: Perspectives on purposes, publicness, and good governance. Public Adm Dev 2011, 31:2–14.

40. Ejaz I, Shaikh BT, Rizvi N: NGOs and government partnership for health systems strengthening: A qualitative study presenting viewpoints of government, NGOs and donors in Pakistan. BMC Health Serv Res 2011, 11(1):122.

41. Hushie M: Public-non-governmental organisation partnerships for health: an exploratory study with case studies from recent Ghanaian experience. BMC Public Health 2016, 16(1):963.

42. Bode I, Brandsen T: State–third Sector Partnerships: A short overview of key issues in the debate. Public Manag Rev 2014, 16(8):1055–1066.

43. Health Systems Learning Group Monograph [https://stakeholderhealth.org/pdf/]

44. Church-health system partnership facilitates transitions from hospital to home for urban, low-income African Americans, reducing mortality, utilization, and costs. Service delivery innovation profile. [http://www.innovations.ahrq.gov/content.aspx?id=3354]

45. Cutts T: The Memphis Model: ARHAP theory comes to ground in the congregational health network. Pietermaritzburg: Cluster Publications; 2010.

46. Carter N, Valaitis R, Feather J, Cleghorn L, Lam A: An Environmental Scan of Health and Social System Navigation Services in an Urban Canadian Community. SAGE Open Nurs 2017, 3:2377960816689566.

47. Corrigan P, Sheehan L, Morris S, Larson JE, Torres A, Lara JL, Paniagua D, Mayes JI, Doig S: The impact of a peer navigator program in addressing the health needs of Latinos with serious mental illness. Psychiatr Serv 2018, 69(4):456–461.

48. Maxwell AE, Jo AM, Crespi CM, Sudan M, Bastani R: Peer navigation improves diagnostic follow-up after breast cancer screening among Korean American women: results of a randomized trial. Cancer Causes Control 2010, 21(11):1931–1940.

49. Pesut B, Duggleby W, Warner G, Fassbender K, Antifeau E, Hooper B, Greig M, Sullivan K: Volunteer navigation partnerships: Piloting a compassionate community approach to early palliative care. BMC Palliat Care 2017, 17(1):2.

50. NHS and Social Care Workforce: Meeting Our Needs Now and in the Future. [https://www.kingsfund.org.uk/sites/default/files/field/field_publication_file/perspectives-nhs-social-care-workforce-jul13.pdf]

51. Engaging Health Care Volunteers to Pursue the Triple Aim. [https://www.aha.org/content/17/17engagingvolunteerstripleaim.pdf]

52. The Role of the Volunteer in Improving Patient Experience [https://www.theberylinstitute.org/store/ViewProduct.aspx?id=6373089]

53. Manderson B, McMurray J, Piraino E, Stolee P: Navigation roles support chronically ill older adults through healthcare transitions: a systematic review of the literature. Health Soc Care Community 2012, 20(2):113–127.

54. Lorhan S, Cleghorn L, Fitch M, Pang K, McAndrew A, Applin-Poole J, Ledwell E, Mitchell R, Wright M: Moving the agenda forward for cancer patient navigation: understanding volunteer and peer navigation approaches. J Cancer Educ 2013, 28(1):84–91.

55. Egan M, Anderson S, McTaggart J: Community navigation for stroke survivors and their care partners: description and evaluation. Top Stroke Rehabil 2010, 17(3):183–190.

56. Arksey H, O’Malley L: Scoping studies: towards a methodological framework. International Journal of Social Research Methodology 2005, 8(1):19–32.

57. Nelson ML, McKellar KA, Munce S, Kelloway L, Hans PK, Fortin M, Lyons R, Bayley M: Addressing the Evidence Gap in Stroke Rehabilitation for Complex Patients: A Preliminary Research Agenda. Arch Phys Med Rehabil 2018, 99(6):1232–1241.

58. Pham MT, Rajić A, Greig JD, Sargeant JM, Papadopoulos A, McEwen SA: A scoping review of scoping reviews: advancing the approach and enhancing the consistency. Res Synth Methods 2014, 5(4):371–385.

59. McHugh ML: Interrater reliability: the kappa statistic. Biochem Med (Zagreb) 2012, 22(3):276–282.

60. Polanin JR, Pigott TD, Espelage DL, Grotpeter JK: Best practice guidelines for abstract screening large-evidence systematic reviews and meta-analyses. Res Synth Methods 2019, 10(3):330–342.

61. Zhao J-G: Combination of multiple databases is necessary for a valid systematic review. Int Orthop 2014, 38(12):2639–2639.

62. Paez A: Gray literature: An important resource in systematic reviews. J Evid Based Med 2017, 10(3):233–240.

